# Genetic Underpinnings and Audiological Characteristics in Children with Unilateral Sensorineural Hearing Loss

**DOI:** 10.1101/2022.12.16.22283544

**Authors:** Chen-Yu Lee, Pei-Hsuan Lin, Yu-Ting Chiang, Cheng-Yu Tsai, Shu-Yu Yang, You-Mei Chen, Chao-Hsuan Li, Chun-Yi Lu, Tien-Chen Liu, Chuan-Jen Hsu, Pei-Lung Chen, Jacob Shujui Hsu, Chen-Chi Wu

**Affiliations:** Department of Otolaryngology, National Taiwan University Hospital, Hsinchu Branch, Hsinchu, Taiwan; Department of Otolaryngology, National Taiwan University Hospital, Taipei, Taiwan; Graduate Institute of Medical Genomics and Proteomics, College of Medicine, National Taiwan University, Taipei, Taiwan; Department of Medical Genetics, National Taiwan University Hospital, Taipei, Taiwan; Division of Pediatric Infectious Diseases, Department of Pediatrics, National Taiwan University Hospital, Taipei, Taiwan; College of Medicine, National Taiwan University, Taipei, Taiwan; Department of Otorhinolaryngology, Taichung Tzu Chi Hospital, Buddhist Tzu Chi Medical Foundation, Taichung, Taiwan; Department of Medical Research, National Taiwan University Hospital, Hsinchu Branch, Hsinchu, Taiwan; Hearing and Speech Center, National Taiwan University Hospital, Taipei, Taiwan

**Keywords:** Unilateral sensorineural hearing loss, *GJB2*, *EDNRB*, *PAX3*, next-generation sequencing

## Abstract

**Objective:** Unilateral sensorineural hearing loss is a condition commonly encountered in otolaryngology clinics. However, its molecular pathogenesis remains unclear. This study aimed to investigate the genetic underpinnings of childhood unilateral sensorineural hearing loss and analyze the associated audiological features.

**Study Design:** Retrospective analysis of a prospectively recruited cohort Setting: Tertiary referral center Methods: We enrolled 38 children with unilateral sensorineural hearing loss and performed physical, audiological, imaging, and congenital cytomegalovirus examinations as well as genetic testing using next-generation sequencing targeting 30 deafness genes. The audiological results were compared across different etiologies.

**Results:** Causative genetic variants were identified in eight (21.1%) patients, including five with *GJB2* variants, two with *PAX3* variants, and one with *EDNRB* variant. *GJB2* variants were associated with mild-to-moderate unilateral sensorineural hearing loss in various audiogram configurations, whereas *PAX3* and *EDNRB* variants were associated with profound unilateral sensorineural hearing loss in flat audiogram configurations. In addition, whole genome sequencing and extended next-generation sequencing targeting 213 deafness genes were performed in two multiplex families compatible with autosomal recessive inheritance; yet no definite causative variants were identified. Cochlear nerve deficiency and congenital cytomegalovirus infection were observed in nine and two patients without definite genetic diagnoses.

**Conclusion:** Genetic underpinnings can contribute to approximately 20% of childhood unilateral sensorineural hearing loss, and different genotypes are associated with various audiological features. These findings highlight the utility of genetic examinations in guiding the diagnosis, counseling, and treatment of unilateral sensorineural hearing loss in children.

## Introduction

Unilateral sensorineural hearing loss (USNHL) is an otologic disorder characterized by hearing impairment in one ear, with preserved audiometric thresholds in the contralateral ear ^1^. USNHL occurs in approximately 1 in 1000 live newborns and may increase in prevalence with age as delayed-onset forms ^2-4^. Various degrees of hearing loss ranging from mild to profound hearing impairment and progression in both severity and laterality have been observed in children with USNHL ^5^.The etiology of USNHL in children differs from that of bilateral sensorineural hearing loss (SNHL). Etiological research showed that 40–66% of children with bilateral SNHL is attributed to genetic causes ^6,7^. In contrast, common causes of USNHL include developmental anomalies in the cochlear nerve or inner ear (∼40%) ^5,6^ and congenital cytomegalovirus (cCMV) infection (∼10%) ^8^.

Identifying the molecular basis is crucial for predicting outcomes and enabling personalized medicine in children with SNHL ^9^. Although USNHL has been occasionally reported in patients with Waardenburg syndrome (WS) ^10^ and some types of non-syndromic SNHI (such as variants in the *GJB2, GJB3, TECTA*, and *COCH* genes) ^11^, the genetic underpinnings have not been comprehensively investigated in children with USNHL. The lack of knowledge regarding molecular etiology hinders the consultation and prognosis of specific treatments for USNHL in children, such as cochlear implantation (CI) ^12^. To address these difficulties, this study aimed to elucidate the genetic underpinnings of USNHL using next-generation sequencing (NGS) and further correlate audiological manifestations with genotypes to facilitate clinical management.

## Methods

### Research Ethics and Patient Consent

This study was conducted in accordance with the principles of the Declaration of Helsinki. This study was approved by the Research Ethics Committee of the National Taiwan University Hospital (IRB number:201803092RINB). All the participants or their parents provided written informed consent.

### Patient Recruitment

We included children (<18 years) who underwent NGS-based genetic examinations for USNHL at National Taiwan University Hospital between January 1, 2018, and December 31, 2021. Diagnoses of USNHL were made by experienced otologists based on audiological data and medical records of each patient. Patients with conductive or mixed-type hearing impairment or those lacking complete medical records were excluded from this study.

### Audiological Tests

Pure tone audiometry (PTA) or diagnostic auditory brainstem response (ABR) was used in audiological assessments depending on the age and cognitive status of the patients ^13^. The first available audiometric test results, either of PTA or ABR, were ascertained to determine the hearing level by averaging the hearing thresholds of 0.5, 1, 2, and 4 kHz. USNHL was defined as a 4-tone hearing level of 25 dB or more in the worse ear and a 4-tone hearing level of less than 25 dB in the contralateral ear ^14^. Furthermore, the severity of SNHL was categorized as mild (25–40 dB), moderate (41–70 dB), severe (71–95 dB), or profound (>95 dB) according to the patient’s 4-tone hearing level. Audiogram configurations were classified as flat, sloping, high-tone hearing loss, or others ^15^.

### Genetic Examination

Peripheral blood samples were obtained from the participants to extract genomic DNA from mononuclear cells, followed by an NGS-based genetic examination targeting 30 common deafness genes ^16^. Through shared decision-making for family study design, the multiplex family USNHL-12 underwent whole genome sequencing (WGS), and the multiplex family USNHL-13 received an extended NGS diagnostic encompassing 213 deafness genes (see **supplemental information S1**). Paired-end reads from the sequencing were aligned, sorted, and detected for single-nucleotide variants (SNVs) and small insertions and deletions (Indels) using Sentieon DNAseq version 2018 ^17^. The identified variants were further annotated using ANNOVAR version 2019 ^18^ and classified according to the 2015 the American College of Medical Genetics and Genomics (ACMG) criteria ^19^. Variants meeting the criteria of pathogenic or likely pathogenic variants were recorded as possible disease-causing variants and confirmed through Sanger sequencing. Finally, a genetic diagnosis was made for participants harboring pathogenic or likely pathogenic variants if their genotypes were compatible with the inheritance mode of the affected gene described in Online Mendelian Inheritance in Man (OMIM) ^20^. A complete list of the equipment, gene panel, and bioinformatics analysis methods applied in this study is shown in **supplemental information S1**.

### Imaging Studies

High-resolution computed tomography (HRCT) of the temporal bone or magnetic resonance imaging (MRI) of the brains were performed for the imaging examination. The decision to obtain images and the choice of imaging modality were individualized per patient and made by the shared decision-making process. A pediatric neuroradiologist and an otologist (C.-C.W.) interpreted the images according to the morphological features of the cochlea, vestibule, semicircular canal, vestibular aqueduct, cochlear aperture, and cochlear nerve. Cochlear nerve deficiency (CND) is defined as the absence or reduction in the caliber of the cochlear nerve compared with the facial nerve ^9^.

### Congenital CMV Testing

Urine CMV isolation tests were performed to confirm cCMV infection. Candidates for the test were those younger than 3 months of age with suspicion of infection after being evaluated by their pediatricians or otolaryngologists.

### Statistical Analysis

Statistical analyses were performed using the R software (version 4.0.5). Categorical data were summarized as numbers (percentages) and analyzed using Fisher’s exact test. Continuous data were analyzed using the Wilcoxon rank-sum test. Statistical significance was set at P < 0.05 (2-sided).

## Results Demographic Data

A total of 38 patients (36 families) with USNHL were included in this study (**Table 1**). The genetic diagnosis was achieved in eight patients (8/38, 21.1%). The mean [standard deviation] age of the patients who were genetically diagnosed (GD) with UHSNL was 3.4 [3.5] years, while that of those who were genetically undiagnosed (GUD) was 3.8 [4.5] years. No differences in age and sex distributions were observed between patients in the GD and GUD groups. Furthermore, no statistical difference in laterality and hearing severity was observed between patients in the GD and GUD groups. The audiological data for each patient are detailed in **Supplemental Table S1**.

**Table 1.**
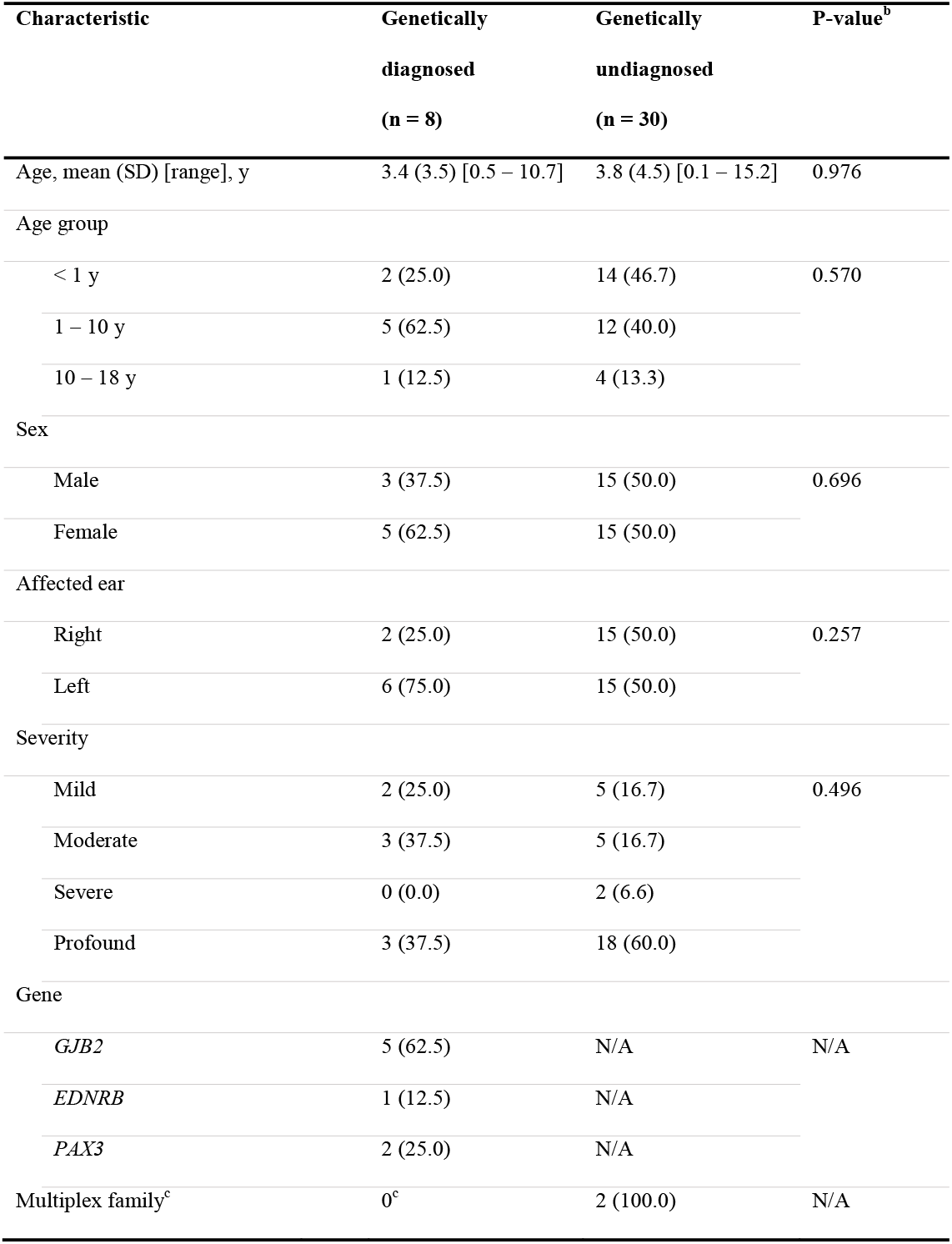
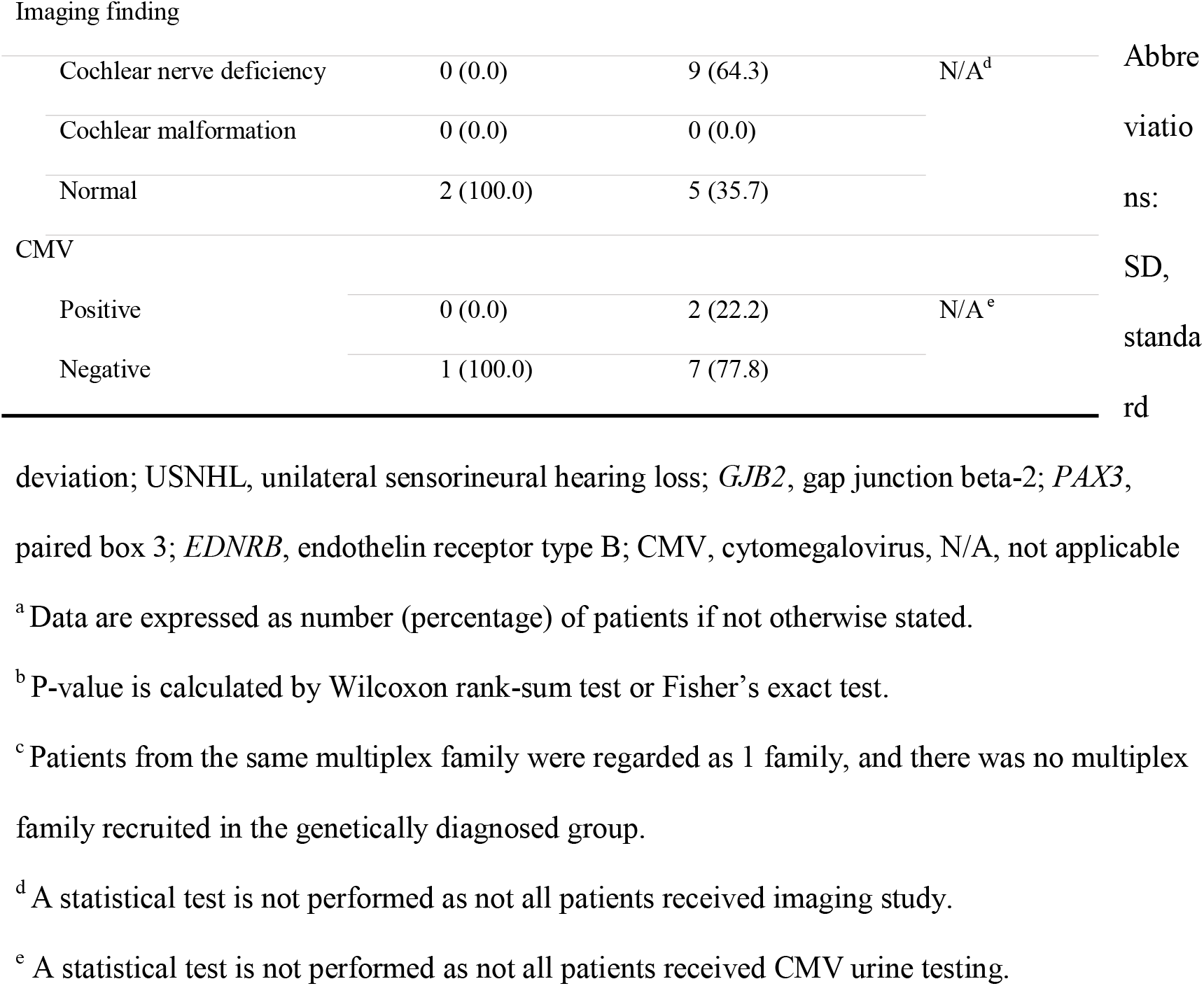
Demographic Characteristics of 38 Patients with USNHL^a^

Sixteen patients (16/38 = 42.1%) underwent imaging studies (2 in the GD group and 14 in the GUD group). CND was identified in 9 (64.3%) of the 14 patients in the GUD group; however, no other types of inner ear malformations were detected in this cohort. CMV tests were performed on 10 patients. Among them, 2 patients who were GUD (2/10, 20%) with profound hearing loss (USNHL-16 and USNHL-21) showed positive urine CMV cultures. Moreover, USNHL-16 had CND, whereas USNHL-21 did not (**Table 1** and **Supplemental Table S1**). The etiology of USNHL remained indefinite in 19 patients (19/38, 50.0%), regardless of comprehensive genetic, imaging, and CMV workups. The distribution of etiological causes in the 38 patients is illustrated in **Supplemental Figure S1**.

### Genotypes and Phenotypic Features of the Patients in the GD Group (n = 8)

Causative genetic variants were identified in eight patients in the GD group, including five (5/8 = 62.5%) with *GJB2* (OMIM 121011) variants, two (2/8 = 25.0%) with *PAX3* (OMIM 606597) variants, and one (1/8 = 12.5%) with *EDNRB* (OMIM 131244) variant (**Table 1**). Four patients were homozygous for the *GJB2* NM_004004.6:c.109G>A (p.Val37Ile) variant, and one had compound heterozygous NM_004004.6:c.[109G>A];[235del] variants. Three patients were GD with WS and distinct heterozygous variants ^21^. USNHL-6-II-1 had a splice site *EDNRB* variant, NC_000013.10(NM_001201397.1):c.754-2A>G, and exhibited heterochromia iridis in addition to hearing impairment. USNHL-7 and USNHL-8, who carried *PAX3* variant NC_000002.12(NM_013942.4): c.587-2A>G and c.1130C>G (p.Ser377Cys), respectively, presented with hearing impairment without additional WS-specific manifestations (**Table 2**).

**Table 2.** Causative Variants Identified in the 8 Patients with USNHL

| Subjects | Variant profile |  |  |  |  | ACMG criteria | Severity |
| --- | --- | --- | --- | --- | --- | --- | --- |
|  | Gene | Variants | Structure | Protein change | Zygosity<sup>a</sup> |  |  |
| USNHL-1 | *GJB2* | NM_004004.6: c.109G>A, Missense | Exon | p.Val37Ile | 1/1 | PP5,PM1,PM5, PS1, PP2 (P) | Mild |
| USNHL-2 | *GJB2* | NM_004004.6: c.109G>A, Missense | Exon | p.Val37Ile | 1/1 | PP5,PM1.PM5,PS1, PP2 (P) | Mild |
| USNHL-3 | *GJB2* | NM_004004.6: c.109G>A, Missense | Exon | p.Val37Ile | 1/1 | PP5,PM1.PM5,PS1, PP2 (P) | Moderate |
| USNHL-4 | *GJB2* | NM_004004.6: c.109G>A, Missense | Exon | p.Val37Ile | 1/1 | PP5,PM1.PM5,PS1, PP2 (P) | Moderate |
| USNHL-5 | *GJB2* | NM_004004.6: c.109G>A, Missense<br>NM_004004.6: c.235del, Frameshift | Exon<br>Exon | p.Val37Ile<br>p.Leu79CysfsTer3 | Compound heterozygous | PP5,PM1.PM5,PS1, PP2 (P, GJB2:c.G109A) PS3, PVS1,PP5,PM2 (P, GJB2:c.235del) | Moderate |
| USNHL-6-II-1 | *EDNRB* | NC_000013.10(NM_001201397.1):c.754-2A>G, Splice site | Intron | N/A | 0/1 | PVS1, PM2 (P) | Profound |
| USNHL-7 | *PAX3* | NC_000002.12(NM_013942.4): c.587-2A>G, Splice site | Intron | N/A | 0/1 | PVS1, PM2 (LP) | Profound |
| USNHL-8 | *PAX3* | NM_181459.4: c.1130C>G, Missense | Exon | p.Ser377Cys | 0/1 | PM2, PP2, PP3 (LP) | Profound |

Abbreviations: USNHL, unilateral hearing loss; *GJB2*, gap junction beta-2; *EDNRB*, endothelin receptor type B; *PAX3*, paired box 3; MAF, minor allele frequency; TB, Taiwan Biobank; ACMG, American College of Medical Genetics and Genomics; N/A, not applicable or not available; P, pathogenic; LP, likely pathogenic
<sup>a</sup> 0/1: heterozygous; 1/1: alternative homozygous

All patients who were GD with *GJB2* variants presented with either mild or moderate USNHL, while those with variants related to WS showed profound USNHL with scaled-out thresholds (**Figure 1**). *GJB2* variants were associated with different audiogram configurations, whereas variants related to WS were associated with a flat audiogram configuration. Noticeably, the mother of USNHL-6-II-1 was identified to have the same pathogenic *EDNRB*:c.754-2A>G variant; however, she did not express any WS symptoms (**Figure 2a & 2b**), suggesting a variant with incomplete penetrance and expressivity. MRI of USNHL-6_II-1 revealed normal cochlear nerves and cochlea in the affected ear (**Figure 2c & 2d**).

**Figure. 1.**
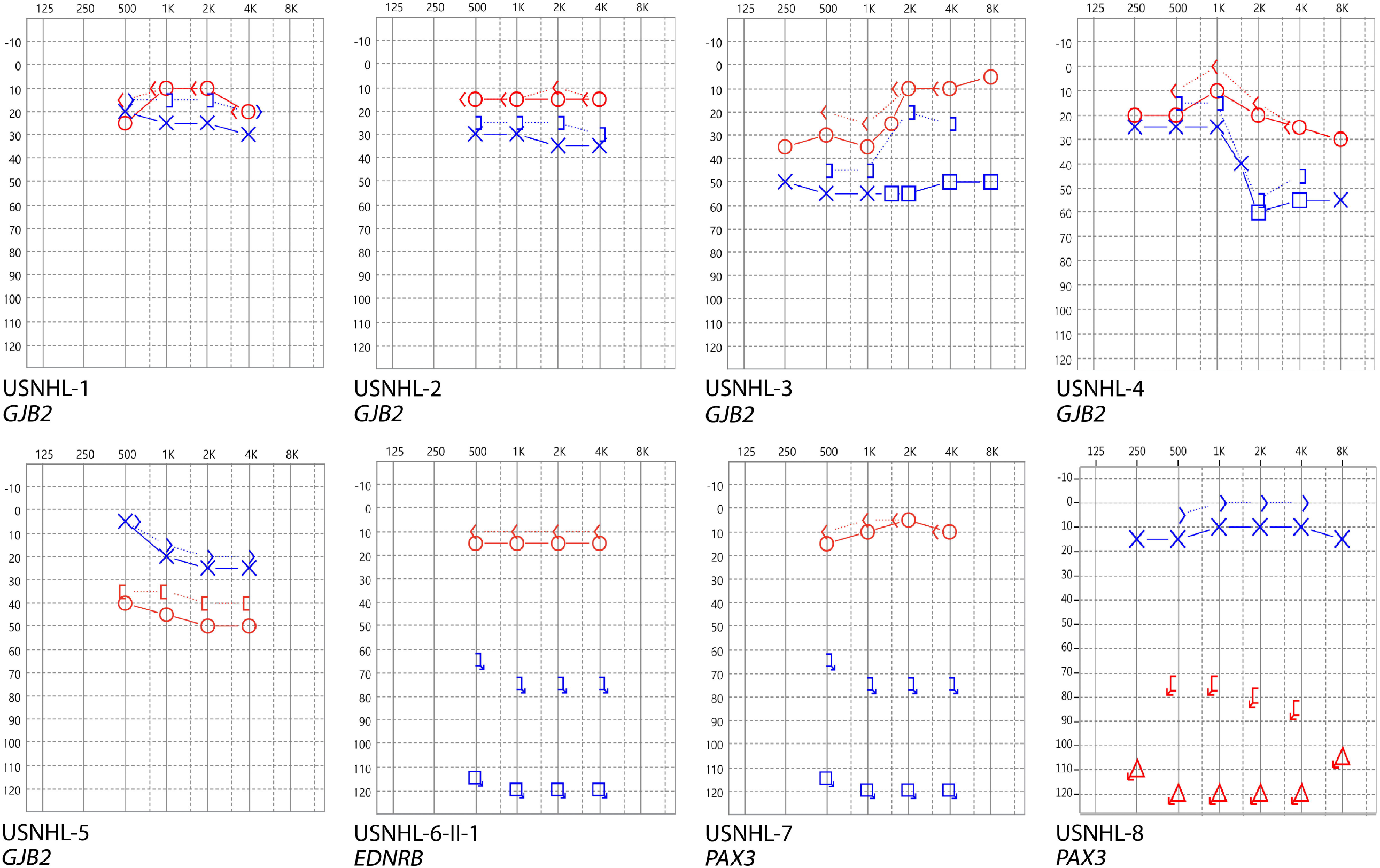
The audiograms of the eight patients who were genetically diagnosed with USNHL. The audiograms of patients: USNHL-1, USNHL-2, USNHL-6-II-1, and USNHL-7 were transformed from their auditory brainstem responses. All the audiograms were used for patients’ USNHL diagnosis. Abbreviations: USNHL, unilateral sensorineural hearing loss; *GJB2*, gap junction beta-2; *EDNRB*, endothelin receptor type B; *PAX3*, paired box 3

**Figure. 2.**
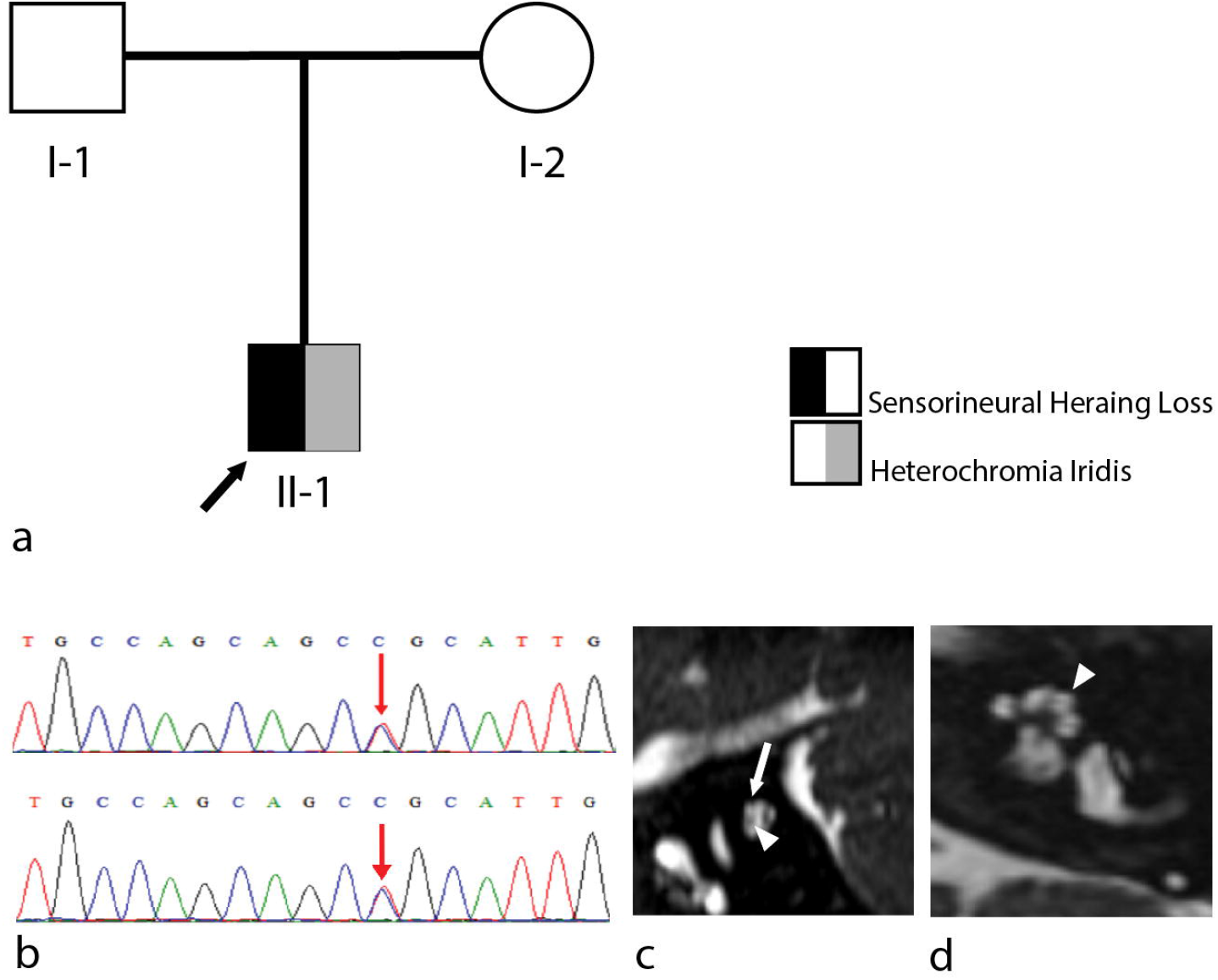
The pedigree, Sanger sequence, and MRI of USNHL-6-II-1 with Waardenburg syndrome. a, USNHL-6-II-1 has sensorineural hearing loss and heterochromia iridis and mother does not express any phenotypes of Waardenburg syndrome while harboring the same variants as he does. b, Sanger sequencing results of the patient (upper sequence) and his mother (lower sequence). c, Oblique sagittal view of the MRI shows a normal-sized left cochlear nerve (arrowhead) compared with the facial nerve (white arrow). d, Axial view of the MRI shows normal cochlear anatomy in USNHL-6-II-1 (arrowhead). Abbreviations: USNHL, unilateral sensorineural hearing loss; MRI, magnetic resonance imaging

### Two multiplex families presenting USNHL

In the family USNHL-12, dizygotic twins II-2 and II-3 exhibited profound hearing impairment on the right side with a flat audiogram configuration (**Figure 3**). In the family USNHL-13, II-1 and II-2 showed moderate hearing impairment with different audiogram configurations ^22^. No progression in severity or laterality and changes in audiogram configurations were noted during the follow-up period in all four patients. No causative variants were identified through WGS in the family USNHL-12 and targeted NGS of 213 deafness genes in the family USNHL-13.

**Figure. 3.**
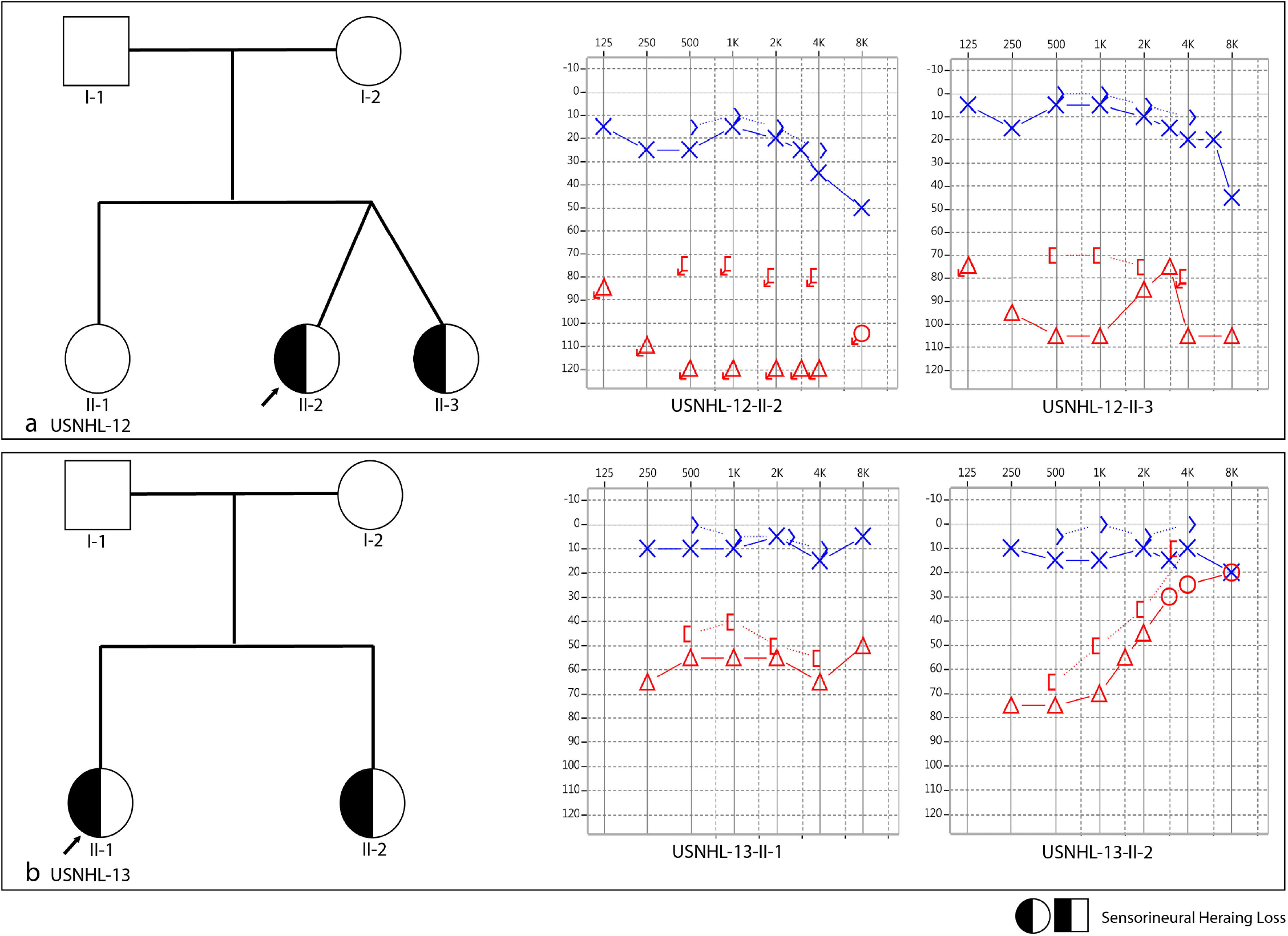
Pedigrees and audiograms of the two multiplex families.

## Discussion

In this study, we achieved genetic diagnoses in 8 (21.1%) of the 38 children with USNHL. Five causative variants of *GJB2, EDNRB*, and *PAX3* were detected, and different genotypes were associated with distinct audiological features. In addition, two multiplex families compatible with autosomal recessive inheritance were speculated to have genetic causes despite lacking a definite genetic diagnosis. We also demonstrated that imaging studies and CMV testing could help in identifying the etiology of USNHL in patients without a definite genetic diagnosis.

*GJB2* encodes connexin26 (Cx26), which acts as the building block of gap junctions and plays an essential role in homeostasis in the organ of Corti ^23^. Defects in Cx26 may interfere with the normal structure and functional maturation of the cochlea, which may lead to hearing loss in mice ^23,24^. In humans, recessive variants of *GJB2* are the most common causes of non-syndromic SNHL ^25^. Among these, *GJB2*:c.109G>A and c.235delC are the two predominant variants in the East Asian population ^7^. *GJB2*:c.109G>A is associated with mild to moderate SNHL with incomplete penetrance, whereas c.235delC is associated with severe-to-profound SNHL ^26,27^. Noticeably, USNHL occurred in 4% of the c.109G>A homozygotes and 5% of the *GJB2*:c.[109G>A];[235del] compound heterozygotes ^26^. Moreover, it has been reported that patients with USNHL with either c.109G>A homozygosity or c.[109G>A];[235del] compound heterozygosity might progress to more severe or bilateral forms of USNHL ^26,28^.

In this study, we identified four patients with USNHL with *GJB2*:c.109G>A homozygosity and one with *GJB2*:c.[109G>A];[235del] compound heterozygosity (**Table 2**). All five patients retained unilateral mild-to-moderate SNHL during at least 6 months of follow-ups. Previous studies have shown that bilateral SNHL caused by homozygous c.109G>A may exhibit diverse ages of onset and affect different frequency ranges ^26,29^. Considering the progressive nature of SNHL associated with c.109G>A, longer follow-up periods might be required to confirm that the unilaterality of SNHL persisted in our five patients. Nonetheless, our results demonstrate that the genetic contribution of *GJB2* variants should not be overlooked in clinical practice, particularly in children with mild-to-moderate USHNL.

WS is a genetic disorder characterized by auditory and pigmentary abnormalities ^30^. Six genes have been confirmed to be responsible for WS: *PAX3* ^31^, *MITF* (OMIM*156845) ^32^, *SOX10* (OMIM*602229) ^33^, *EDN3* (OMIM*131242) ^34^, *EDNRB* ^35^, and *SNAI2* (OMIM*602150) ^36^.

Defects in the WS genes were found to affect the differentiation and development of neural crest cells (NCCs) into intermediate cells (ICs) in the stria vascularis (SV) ^37,38^. Abnormalities in ICs lead to aberrant homeostasis of potassium ions in the endolymph, which may dysregulate the endolymphatic environment, leading to hair cell apoptosis and eventually SNHL ^39-41^. USNHL has been reported in several WS studies ^42,43^. Song et al. reported in a meta-analysis study that 10.6% of patients with WS had SNHL were unilateral and demonstrated that USNHL in WS might be gene-dependent; USNHL was presented in 20.0–28.6% of patients with WS with *PAX3, EDNRB*, or *END3* variants and was shown in only 0–10.5% of patients with WS with *MITF, SOX10*, or *SNAI2* variants ^44^. Furthermore, genetic background and modifiers were suspected to be involved in the penetrance of SNHL in WS, contributing to diverse audiologic phenotypes, including USNHL ^43^.

In this study, two patients harbored likely pathogenic variants in *PAX3* and one harbored a pathogenic *EDNRB* variant. All three patients with WS presented with scaled-out USNHL (**Figure 1**). These findings were consistent with those of previous reports that while the severity of WS in patients varied within and between families, profound SNHL was the most common audiological phenotype ^37^.

Notably, both multiplex families (USNHL-12 and 13) with USNHL showed a recessive mode of inheritance (**Figure 3**). Although no disease-causing SNVs or Indels were identified through our WGS and extended targeted NGS in these two families, the genetic contribution to USNHL could not be excluded. USNHL is believed to affect approximately 1 in 1,000 newborns congenitally ^45^. Therefore, the hereditary roots of USNHL must be statistically justified if more than one case is found in the same family ^46^. Given the limitations of the current NGS-based diagnostic platforms ^47^, the genetic underpinnings of the two families could be structural variants, pathogenic variants in novel deafness genes, or disease-causing intronic variants.

For patients without a definite genetic diagnosis in this study, CND accounted for 64.3% of their etiologies, and cCMV infection was responsible for a small proportion of USNHL cases (**Supplemental Figure S1**). Our results are compatible with previous findings that CND and inner ear malformations are common causes of USNHL ^48^. Nevertheless, there may still be genetic determinants that underlie these two origins. For example, CND has been reported in patients with CHARGE, VACTERL, and Coffin-Siris syndromes ^49-51^, which are hereditary and associated with USNHL ^52^. Furthermore, multiple genes have been documented involved in inner ear malformations in syndromic and non-syndromic SNHL ^53,54^. Therefore, considering the aggregation of patients with USNHL in multiplex families and the potential association between USNHL and genetics-related inner ear malformations, it is inferred that the genetic component of USNHL may be higher than the 21.1% demonstrated in this study.

The management of USNHL in children who cannot attain binaural hearing using conventional hearing aids remains controversial. CI has shown benefits in restoring binaural hearing in children with profound USNHL ^55,56^; however, the Food and Drug Administration has only approved CI in patients aged 5 years or older ^12^. One of the concerns regarding CI in prelingual children with USNHL arises from its uncertain outcomes. Structural anomalies, such as CND and inner ear malformations, which are common etiologies of USNHL in children, may lead to limited benefits with CI ^57^. In contrast, patients with prelingual WS with profound USNHL may be good candidates for CI. First, as the main pathogenetic mechanism of WS is NCC dysfunction in the stria vascularis, and the pathology is confined to the inner ear, the cochlear nerve and cochlea are intact for CI stimulation, as demonstrated in the imaging study of our USNHL-6-II-1 patient. Furthermore, except for *SOX10* variants, patients with most types of WS do not exhibit inner ear malformations ^44^. As such, favorable CI outcomes have been reported in patients with WS with bilateral SNHL ^58^. It is conceivable that with the same pathogenetic mechanisms, favorable CI outcomes can be anticipated in patients with WS with USNHL. In contrast, patients with *GJB2* variants may only require observation or hearing aids because of the slow progression and mild-to-moderate severity of SNHL ^28^. Taken together, the identification of genetic causes may assist in precision medicine by tailoring clinical assessment and treatment protocols for children with USNHL.

In this study, we demonstrated the utility of genetic examination in delineating the molecular causes of USNHL and clarified specific genotype-phenotype correlations that can facilitate the assessment, counseling, and management of children with USNHL in clinical practice. However, this study has several limitations that deserve further discussion. First, the NGS diagnostics adopted in this study only screened for exonic and splice site variants in 30 common deafness genes. Therefore, pathogenic variants of other uncommon deafness genes were not addressed. Similarly, copy number variations (CNVs) ^59^ and variants in non-coding regions ^60^ were beyond the scope of the NGS diagnostics used in this study. Second, the generalizability of this study could be limited due to the relatively small cohort size of 38 patients from a single Han Taiwanese population. Nonetheless, the diagnostics and cohort size limitations did not compromise our findings regarding the role of genetic factors in childhood USNHL.

In conclusion, we confirmed that genetic underpinnings could contribute to approximately 20% of childhood USNHL and demonstrated that different genotypes were associated with different audiologic features and outcomes. These findings indicate the potential utility of including genetic examination in the evaluation battery for childhood USNHL, as the identification of the genetic etiology can help guide appropriate counseling and management of the disease.

## Data Availability

All data produced in the present study are available upon reasonable request to the authors.

## Acknowledgments

We thank all the patients and their relatives participating in this study. The high-coverage targeted next-generation sequencing data was generated at the Laboratory of Molecular Genetic Diagnostics at the National Taiwan University Hospital. We also thank National Center for High-performance Computing (NCHC) for providing computational and storage resources.

## Data Availability

All data produced in the present study are available upon reasonable request to the authors.

## Figure and Supplemental Legends

**Supplemental Figure. 1** Pie chart illustrating the etiologies of the 38 patients with USNHL. Abbreviations: cCMV, congenital cytomegalovirus infection; USNHL, unilateral sensorineural hearing loss

