## Supplementary figures and images for "Genetic Underpinnings and Audiological Characteristics in Children with Unilateral Sensorineural Hearing Loss"

### Supplemental Figure S1

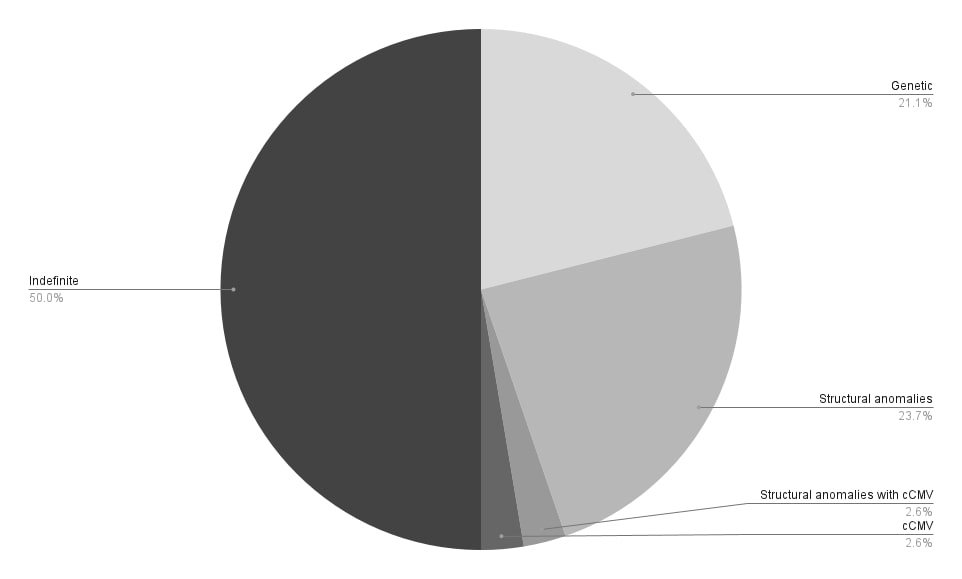
