## Supplemental Information S1 for "Genetic Underpinnings and Audiological Characteristics in Children with Unilateral Sensorineural Hearing Loss"

**Genetic Examination**

**Next-Generation Sequencing**

After the participants’ mononuclear cells were collected, a sonication method (Covaris, Woburn, MA, USA) was used to produce DNA fragments with an average size of 800 bp. A 2100 Bioanalyzer (Agilent Technologies, Santa Clara, CA, USA) was used to measure the length of the fragments generated, and Qubit (Thermo Scientific, Waltham, MA) was used to measure fragment concentration.  A DNA library was constructed from the fragments using a TruSeq Library Preparation Kit (Illumina Inc., San Diego, CA, USA). In this study, the SeqCap EZ Hybridization and Wash Kit (Roche NimbleGen, Madison, WI) was used to achieve capture-based target enrichment by employing probes designed to target the coding regions of 30 common deafness-associated genes in the Taiwanese population (see “30 Common Deafness-Associated Genes Panel” below). The total size of the target region was approximately 317 kb. Finally, all DNA samples were sequenced using the MiSeq platform (Illumina Inc., San Diego, CA) to produce 300-nucleotide paired-end reads with a 150x average read depth. The affected proband and her mother from the USNHL-12 family underwent whole genome sequencing (WGS) using the NovaSeq 6000 System (Illumina Inc., San Diego, CA) with 150bp paired-end fragments [1]. Another two patients from the USNHL-13 family underwent targeted gene sequencing using the same platforms and sample preparation methods mentioned above, yet with an extended 213 deafness-associated gene panel for genomic sequencing (see “Extended 213 Deafness-Associated Genes Panel” below).

**Variant Analyses**

Sequence alignment, sorting, and file conversion were performed using the paired-end reads from the next-generation sequencing with the BWA-MEM and Sort utility in the Sentieon DNAseq version 2018 (<https://www.sentieon.com/products/>) [2]. Single nucleotide substitutions and small insertions and deletions were detected through the Sention Haplotyper algorithm. All identified variants from the previous steps were further annotated by ANNOVAR version 2019 ([https://annovar.openbioinformatics.org/en/latest/#annovar-documentation](https://annovar.openbioinformatics.org/en/latest/" \l "annovar-documentation)) [3]. A series of information was annotated to the variants, including Human Genome Variation Society nomenclatures, maximum allele frequency across distinguished populations of the gnomAD database [4], minor allele frequency in the Taiwan Biobank database [5], and various *in silico* prediction outcomes, including PolyPhen-2 version 2 (Harvard University, Cambridge, MA), SIFT version 2019 (SANS Institute, North Bethesda, MD), LRT version 2009 (Washington University, St. Louis, MO), MutationTaster version 2 (Charité e Universitätsmedizin Berlin, Berlin, Germany), VariantAssessor version 3 (Computational Biology Center, Memorial Sloan-Kettering Cancer Center, NY), FATHMM version 2.3 (University of Bristol, Bristol, England), and MetaLR version 2015 (Human Genetics Center, University of Texas Health Science Center at Houston, Houston, TX).

Variants located at exomes or intron splicing sites were selected as candidate variants. The online platform VarSome [6] was adopted to carefully assess all candidate variants in accordance with the American College of Medical Genetics and Genomics (ACMG) guidelines [7]. Variants meeting the criteria of pathogenic and likely pathogenic variants were recorded as possible disease-causing and confirmed through Sanger sequencing.

| **30 Common Deafness-Associated Genes Panel** | | | | | | | |
| --- | --- | --- | --- | --- | --- | --- | --- |
| *AIFM1* | *DFNB59* | *DIAPH3* | *EDN3* | *EDNRB* | *EYA1* | *FOXI1* | *GJA1* |
| *GJB1* | *GJB2* | *GJB3* | *GJB4* | *GJB6* | *KCNJ10* | *KCNQ4* | *MITF* |
| *PAX3* | *PCDH9* | *MTRNR1* | *MYO15A* | *OTOF* | *POU3F4* | *POU4F3* | *SIX5* |
| *SIX1* | *SLC26A4* | *SNAI2* | *STRC* | *SOX10* | *TMPRSS3* |  |  |

| **Extended 213 Deafness-Associated Genes Panel** | | | | | | | |
| --- | --- | --- | --- | --- | --- | --- | --- |
| *ACTB* | *ACTG1* | *ADCY1* | *ADGRV1* | *AIFM1* | *ALMS1* | *ATP2B2* | *ATP5MF* |
| *ATP6V0A4* | *ATP6V1B1* | *ATP6V1B2* | *BCS1L* | *BDP1* | *BSND* | *BTD* | *CA2* |
| *CABP2* | *CACNA1D* | *CATSPER2* | *CCDC50* | *CD164* | *CDC14A* | *CDH23* | *CEACAM16* |
| *CEMIP* | *CEP78* | *CHD7* | *CIB2* | *CISD2* | *CLDN14* | *CLIC5* | *CLPP* |
| *CLRN1* | *COCH* | *COL11A1* | *COL11A2* | *COL2A1* | *COL4A3* | *COL4A4* | *COL4A5* |
| *COL4A6* | *COL9A1* | *COL9A2* | *COL9A3* | *CRYL1* | *CRYM* | *DCDC2* | *DIABLO* |
| *DIAPH1* | *DIAPH3* | *DMXL2* | *DSPP* | *DUOX2* | *ECE1* | *EDN3* | *EDNRA* |
| *EDNRB* | *ELMOD3* | *EPS8* | *EPS8L2* | *ERAL1* | *ERCC2* | *ERCC3* | *ESPN* |
| *ESRRB* | *EYA1* | *EYA4* | *FAS* | *FGF3* | *FGFR1* | *FGFR2* | *FGFR3* |
| *FITM2* | *FOXI1* | *GATA3* | *GIPC3* | *GJA1* | *GJB1* | *GJB2* | *GJB3* |
| *GJB4* | *GJB6* | *GPSM2* | *GRHL2* | *GRXCR1* | *GRXCR2* | *GSDME* | *GSTP1* |
| *HAL* | *HARS* | *HARS2* | *HECTD3* | *HGF* | *HOMER2* | *HSD17B4* | *ILDR1* |
| *JAG1* | *KARS1* | *KCNE1* | *KCNJ10* | *KCNJ11* | *KCNQ1* | *KCNQ4* | *KITLG* |
| *LARS2* | *LHFPL5* | *LHX3* | *LOXHD1* | *LOXL3* | *LRP5* | *LRTOMT* | *MARVELD2* |
| *MCM2* | *MET* | *MIR182* | *MIR183* | *MIR96* | *MITF* | *MPZ* | *MSRB3* |
| *MTAP* | *MYH14* | *MYH9* | *MYO15A* | *MYO1C* | *MYO1F* | *MYO3A* | *MYO6* |
| *MYO7A* | *NARS2* | *NDP* | *NF2* | *NLRP3* | *NR2F1* | *OPA1* | *OSBPL2* |
| *OTOA* | *OTOF* | *OTOG* | *OTOGL* | *OTOR* | *P2RX2* | *PAX3* | *PCDH15* |
| *PCDH9* | *PDZD7* | *PEX1* | *PEX6* | *PEX7* | *PHYH* | *PJVK* | *PMP22* |
| *PNPT1* | *POLR1C* | *POLR1D* | *POU3F4* | *POU4F3* | *PRPS1* | *PTPRQ* | *PTRH2* |
| *RDX* | *RIPOR2* | *ROR1* | *S1PR2* | *SCARB2* | *SEMA3E* | *SERPINB6* | *SIX1* |
| *SIX5* | *SLC17A8* | *SLC22A4* | *SLC26A4* | *SLC26A5* | *SLC4A1* | *SLC4A11* | *SLC52A2* |
| *SLC52A3* | *SLC6A13* | *SLITRK6* | *SMPX* | *SNAI2* | *SOX10* | *SOX2* | *SPINK5* |
| *STRC* | *SYNE4* | *TBC1D24* | *TBL1X* | *TBX1* | *TCF21* | *TCOF1* | *TECTA* |
| *TECTB* | *TFCP2* | *TIMM8A* | *TJP2* | *TMC1* | *TMEM126A* | *TMEM132E* | *TMIE* |
| *TMPRSS3* | *TMPRSS5* | *TNC* | *TPRN* | *TRIOBP* | *TSPEAR* | *TWNK* | *USH1C* |
| *USH1G* | *USH2A* | *WBP2* | *WFS1* | *WHRN* |  |  |  |
