## Supplemental Table S1 for "Genetic Underpinnings and Audiological Characteristics in Children with Unilateral Sensorineural Hearing Loss"

| **Supplemental Table S1 Audiological, Genotype, Imaging, and CMV Testing Results of 38 Patients with USNHL** | | | | | | | | |
| --- | --- | --- | --- | --- | --- | --- | --- | --- |
| Subjects | Age range, y | Sex | Affected side | Severity | Variant identified | Zygosity^a^ | Imaging finding | CMV^b^  testing |
| USNHL-1 | 0-5 | M | Left | Mild | *GJB2*(NM_004004.6):c.G109A (p.Val37Ile), Missense | 1/1 | N/A | N/A |
| USNHL-2 | 0-5 | F | Left | Mild | *GJB2*(NM_004004.6):c.G109A (p.Val37Ile), Missense | 1/1 | N/A | N/A |
| USNHL-3 | 6-10 | M | Left | Moderate | *GJB2*(NM_004004.6):c.G109A (p.Val37Ile), Missense | 1/1 | Normal | N/A |
| USNHL-4 | 0-5 | F | Left | Moderate | *GJB2*(NM_004004.6):c.G109A (p.Val37Ile), Missense | 1/1 | N/A | N/A |
| USNHL-5 | 0-5 | F | Right | Moderate | *GJB2*(NM_004004.6):c.G109A (p.Val37Ile), Missense *GJB2* (NM_004004.6):c.235del (p.Leu79CysfsTer3), Frameshift | Compound heterozygous | N/A | N/A |
| USNHL-6-II-1 | 0-5 | M | Left | Profound | *EDNRB*(NM_001201397.1):c.7542A>G, Splice site | 0/1 | N/A | N/A |
| USNHL-7 | 0-5 | F | Left | Profound | *PAX3*(NM_013942.5):c.5872A>G, Splice site | 0/1 | N/A | Negative |
| USNHL-8 | 6-10 | F | Right | Profound | *PAX3*(NM_181459.4):c.1130C>G , Missense | 0/1 | N/A | N/A |
| USNHL-9 | 6-10 | F | Left | Mild | *GJB2*(NM_004004.6):c.G109A (p.Val37Ile), Missense | 0/1 | N/A | N/A |
| USNHL-10 | 0-5 | M | Left | Mild | *MYO15A*(NM_016239.4):c.6863C>T (p.Ser2288Leu), Missense | 0/1 | N/A | N/A |
| USNHL-11 | 0-5 | M | Right | Profound | *OTOF*(NM_194248.3):c.5098G>C (p.Glu1700Gln), Missense | 0/1 | N/A | N/A |

| **Supplemental Table S1 (continued)** | | | | | | | | |
| --- | --- | --- | --- | --- | --- | --- | --- | --- |
| USNHL-12-II-2 | 11-15 | F | Right | Profound | Not found | N/A | Normal | N/A |
| USNHL-12-II-3 | 11-15 | F | Right | Profound | Not found | N/A | Normal | N/A |
| USNHL-13-II-1 | 6-10 | F | Right | Moderate | Not found | N/A | N/A | N/A |
| USNHL-13-II-2 | 6-10 | F | Right | Moderate | Not found | N/A | N/A | N/A |
| USNHL-14 | 6-10 | F | Left | Profound | Not found | N/A | N/A | N/A |
| USNHL-15 | 0-5 | F | Right | Profound | Not found | N/A | Cochlear nerve deficiency | N/A |
| USNHL-16 | 0-5 | F | Right | Profound | Not found | N/A | Cochlear nerve deficiency | Positive |
| USNHL-17 | 0-5 | F | Left | Profound | Not found | N/A | Cochlear nerve deficiency | N/A |
| USNHL-18 | 0-5 | M | Right | Moderate | Not found | N/A | N/A | N/A |
| USNHL-19 | 0-5 | M | Left | Mild | Not found | N/A | Normal | N/A |
| USNHL-20 | 0-5 | F | Right | Severe | Not found | N/A | N/A | N/A |

| **Supplemental Table S1 (continued)** | | | | | | | | |
| --- | --- | --- | --- | --- | --- | --- | --- | --- |
| USNHL-21 | 0-5 | M | Left | Profound | Not found | N/A | N/A | Positive |
| USNHL-22 | 0-5 | M | Right | Mild | Not found | N/A | N/A | Negative |
| USNHL-23 | 0-5 | F | Left | Profound | Not found | N/A | Normal | N/A |
| USNHL-24 | 0-5 | M | Right | Profound | Not found | N/A | N/A | N/A |
| USNHL-25 | 0-5 | F | Right | Profound | Not found | N/A | N/A | Negative |
| USNHL-26 | 0-5 | F | Right | Profound | Not found | N/A | Cochlear nerve deficiency | Negative |
| USNHL-27 | 0-5 | M | Right | Profound | Not found | N/A | N/A | Negative |
| USNHL-28 | 11-15 | F | Left | Moderate | Not found | N/A | Normal | Negative |
| USNHL-29 | 0-5 | M | Left | Mild | Not found | N/A | N/A | N/A |
| USNHL-30 | 6-10 | M | Left | Severe | Not found | N/A | Cochlear nerve deficiency | N/A |
| USNHL-31 | 0-5 | M | Right | Moderate | Not found | N/A | N/A | N/A |
| USNHL-32 | 0-5 | M | Left | Profound | Not found | N/A | Cochlear nerve deficiency | N/A |
| USNHL-33 | 0-5 | M | Left | Profound | Not found | N/A | Cochlear nerve deficiency | Negative |

| **Supplemental Table S1 (continued)** | | | | | | | | |
| --- | --- | --- | --- | --- | --- | --- | --- | --- |
| USNHL-34 | 0-5 | F | Left | Profound | Not found | N/A | Cochlear nerve deficiency | Negative |
| USNHL-35 | 0-5 | M | Left | Profound | Not found | N/A | N/A | N/A |
| USNHL-36 | 0-5 | M | Left | Profound | Not found | N/A | Cochlear nerve deficiency | N/A |

Abbreviations: USNHL, unilateral sensorineural hearing loss; M, male; F, female; *GJB2*, gap junction beta-2; *PAX3*, paired box 3; *EDNRB*, endothelin receptor type B; *MYO15A*, myosin XVA; *OTO*F, otoferlin; CMV, Cytomegalovirus, N/A, not applicable or not performed

^a^ 0/1: heterozygous; 1/1: alternative homozygous

^b^ Testing is considered as positive if CMV is isolated from a patient’s urine sample.
